# Pre-school Age Participation in Mass Drug Administration: Analysing the Impact on Community-wide Schistosomiasis Control

**DOI:** 10.1101/2025.03.06.25323487

**Authors:** John R. Ellis, Roy M. Anderson

## Abstract

**Background:** Schistosome infection in childhood is common and can lead to morbidity. A formulation of praziquantel to treat pre-school-aged children (PSAC) has been developed recently. This paper assesses what impact including PSAC in mass drug administration (MDA) may have on transmission and morbidity at a community-wide level.

**Methods:** We utilise a model of schistosome transmission to simulate the probability of a community reaching elimination as a public health problem (EPHP) and the reduction in morbidity of children resulting from infections until the age of five, measured by a ‘Worm Years’ metric as a score of morbidity.

**Results:** Including PSAC in MDA will almost always lead to a reduction in morbidity. However, it does not necessarily result in a substantial increase in the probability of EPHP. The proportion of schistosome infections in each age group is a key factor in determining the effectiveness of MDA programmes which prioritise different age groups for treatment.

**Conclusions:** Policy makers should be aware that including PSAC in MDA may not help to reach the WHO target of EPHP. However, a reduction in the average summed worm infection burden at the age children typically start attending school is highly desirable in increasing the long-term benefit of MDA in early childhood.

## Introduction

Schistosomiasis is a parasitic disease, caused by the trematode worms in the genus Schistosoma. It affects more than 230 million people in Africa, Asia, South America and the Caribbean.^1^ Mature worms live in either the intestines (Schistosoma mansoni or S. japonicum) or in the urogenital tract (S. haematobium) where they mate and release eggs.^2^ The World Health Organization (WHO) have set the target for the elimination of schistosomiasis as a public health problem globally by 2030, and the more ambitious target of interruption of transmission in selected countries.^1^ The main method of control is mass drug administration (MDA) employing the drug praziquantel (PZQ), primarily targeting school-age children (SAC).^3^

Previously, pre-school aged children (PSAC) have not been offered treatment due to concerns about the safety, dosage, and method of application.^4^ However, recently there have been calls to include PSAC in MDA, due to recognition of much morbidity in this age group in some settings,^5,6^ and due to progress in determining an optimal safe dosage.^7^ Although it was previously assumed that prevalence among PSAC was low, and therefore contribute little towards transmission, many studies have shown high prevalence in PSAC, particularly in endemic areas with high transmission rates.^8,9^ The WHO now recommends annual preventative chemotherapy for all age groups from two years old in moderate and high prevalence areas and access to treatment to control morbidity for all ages, including PSAC younger than two years of age.^1^

To facilitate this, a new formulation suitable for PSAC, called arpraziquantel (L-PZQ) has been developed by Merck KGaA and the Pediatric Praziquantel Consortium and has recently received a positive scientific opinion from the European Medicines Agency. It has recently been included in the WHO list of prequalified medicines.^10^ L-PZQ is an orodispersible tablet designed to be more child-friendly, without the bitter taste associated with PZQ.^11^ Clinical studies found that this formulation was palatable to PSAC and clinical trials have shown that is has similar efficacies to PZQ in SAC and adults.^11,12^

Morbidity from schistosome infection does not always relate to the current intensity of infection, usually measured by egg counts in faeces or urine.^13^ Severe morbidity is typically related to overall duration of exposure to infection over many years.^4,14^ This is particularly true in the case of PSAC, where the damage induced by infection accumulates. Morbidity in SAC and adults is therefore likely to be related to the history of infection in earlier life as well as by the current infection levels.^14,15^ Some of the effects of infection can be reversed if treated while still a child, if rapid reinfection does not occur.^16^ It follows that the earlier treatment is received, the larger the potential impact it will have on a child’s health. There are also additional adverse effects from infection at young age, for example, chronic exposure is associated with reduced efficacy of childhood vaccines.^6^ Schistosome infection is also associated with declines in school attendance, educational achievement and cognitive deficits.^17^ Clearly, on an individual level, treatment with L-PZQ will benefit PSAC with high burdens of infection and prevent them from entering the school system with associated morbidities.

To determine the value of including PSAC in regular MDA programmes, it is important to consider the impact across the whole population including all age groups. To do this, one must consider both the cross-sectional age intensity profile of infection (i.e. how the rate of infection of a host changes with age), and the demography of the community. It is known that infection peaks during the school-aged years,^18^. In resource poor settings with high net birth rates this group makes up a substantial proportion of the total population. Therefore, MDA in SAC targets a large proportion of infections (POI) in the total population. However, in communities that have received many years of MDA for SAC but have been unsuccessful in reaching elimination, this age distribution may have changed over time with improved child survival.^19^ Mathematical models can be used to simulate the effect of different MDA treatment strategies on the population both in terms of prevailing levels of infection and the accumulated past exposure to infection in defined age groups. Rather than relying on estimates of the burden of infection, the full infection history of a host can be recorded by individual based stochastic models that trac infection exposure and treatment history in all individuals within a community. Individual based models have previously been used, for example, to show the required MDA coverage needed to achieve WHO targets, the impact of non-compliance on control impact and the post-treatment surveillance needed to confirm elimination either as a public health problem or of transmission.^18,20,21^

In this paper, we analyse previously published data to assess the distribution of all infections across ages in the population prior to the implementation of MDA. An individual-based stochastic model is employed to calculate both the effects of including PSAC in MDA programmes on the prevalence of schistosome infection while varying the contact rates of the PSAC population, and the ability to meet WHO target of elimination as a public health problem (EPHP).^2^ We then assess the impact of paediatric drug administration on childhood health using prevalence, infection intensity and a metric based on the total number of infections experienced during an individual’s lifetime (Worm Years) as a proxy for morbidity.

## Methods

### Data

The impact of paediatric PZQ will depend on a number of key epidemiological parameters. How infection intensity varies by the age of the host is of great importance. Typically, the prevalence and mean intensity of infection in a population peaks in SAC.^18^ However, the shape of the age profile varies between locations and can be dependent on geography, climate, access to sanitation and previous exposure to treatment.^6,22^ Another key factor that will determine the impact of treating PSAC is the demography of the population being treated, which will also vary between countries and locations within them.

We compare the age profiles taken from four age and gender stratified large sample size studies; two are S. haematobium epidemiological datasets from Tanzania and Zimbabwe prior to MDA,^23,24^ and two are S. mansoni studies from Kenya prior to mass treatment.^25^ We have used the US census bureau to recover the population pyramid in the respective countries close to the time of each epidemiological survey to calculate the proportion of the total parasite population (POI) that are within each age grouping. ^26^ Where egg count data is not given for a specific age, a simple linear extrapolation method is employed using data for age groups belwo and above the missing data group to fill in the gaps. The POI for each age is calculated as the mean egg count multiplied by the proportion of the population, normalised so the sum over all ages is equal to one.

### Individual based Stochastic Model

The individual based stochastic model of schistosome transmission and control by MDA is described in detail in past publications.^18,27^ To compare the results of MDA with and without the inclusion of PSAC, we consider two approaches. Firstly, we consider whether treating PSAC can improve the chances of meeting the WHO goal of elimination as a public health problem (EPHP), achieved when the prevalence of heavy intensity infection in SAC is reduced to less than 1%.^2^ Heavy intensity infection is defined as ≤ 400 epg for intestinal schistosomiasis and 50 eggs/10 ml for urogenital schistosomiasis.^2^

To assess the impact of MDA on morbidity, we introduce the metric of worm years (WY), which measures infection intensity over the entire lifespan of a host. We utilise this metric to assess the benefits achieved by different MDA strategies, specifically in young ages groups, by calculating the WY at specific ages and the subsequent reduction in the mean WY due to MDA. To calculate a host’s

WY, we take the sum of time spent infected by each worm since birth. This is the same as taking the integral of the worm burden for each individual, *i*, from 0 to their current age (*a_i*):

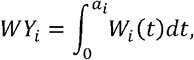

where *W_i*(*t*) is the worm burden of individual *i* at time *t*,. We focus on children between the ages of five and six as that is the age when they start to enter education and may be affected by disease/morbidity induced by the infections they have had up until this point.

We assume that MDA of SAC is prioritised in accord with current WHO guidelines and consider scenarios where coverage is either at 75% or 60% of this age group. We then compare the inclusion of PSAC and adults in the MDA programme at the same coverage level as SAC. MDA is administered annually to the population with a beta binomial model of adherence, so that there is variation in the probability of being treated at each round between individuals in line with published studies of compliance patterns ^28,29^. We assume that the efficacy of L-PZQ for PSAC is the same as PZQ is for other age groups.^11,12^

We restrict our attention to the infection intensity age profile of S. haematobium in Misungwi, Tanzania,^24^ and S. mansoni in Matithini, Kenya,^25^ where past studies have estimated the key epidemiological parameters.^18,20^ The full set of parameters employed in the simulations are listed in Tables S1 and S2 in the supplementary material. However, for the model of S. haematobium in Tanzania, we also change the contact rate of the youngest age group (β_(0-5)_) to vary the proportion of infection in PSAC. The demography of the population is initially fitted to modern day (2024) Tanzania, however this is also varied to explore the effect of past and predicted future demographic trends, by fitting to past (1978) and future (2070) estimates of the demography of Tanzania.^26^ Results are based on 300 simulations of a population of 500 people for each parameter set.

## Results

Mean egg counts, demography and POI in the four datasets are presented in Figure 1, at yearly age intervals and colour coded by age group. Total POI for each age group is also listed in Figure 1, panels I-L. There is a great deal of variation in mean egg counts, particularly between the two S. mansoni datasets in Kenya (Figure 1A and B), where infection intensity is relatively low in PSAC and high in adults, compared to the S. haematobium datasets (Figure 1C and D), where the infection intensity in adults in particular is much lower. This difference in age-intensity profiles is widely recorded for these two infections. The demography is similar for all but the Zimbabwe dataset (Figure 1H), which was collected more recently than the others and due to improved child and teenage survival there is a higher proportion of adults. Despite this, 11.5% of infections are in PSAC, compared to 2.1% and 2.5% in the two Kenya datasets, where the proportion of PSAC in the population is larger. However the greatest POI in PSAC can be found in the Tanzania dataset, where 12% of infections are in PSAC, 60.1% in SAC and only 27.8% in adults.

**Figure 1:**
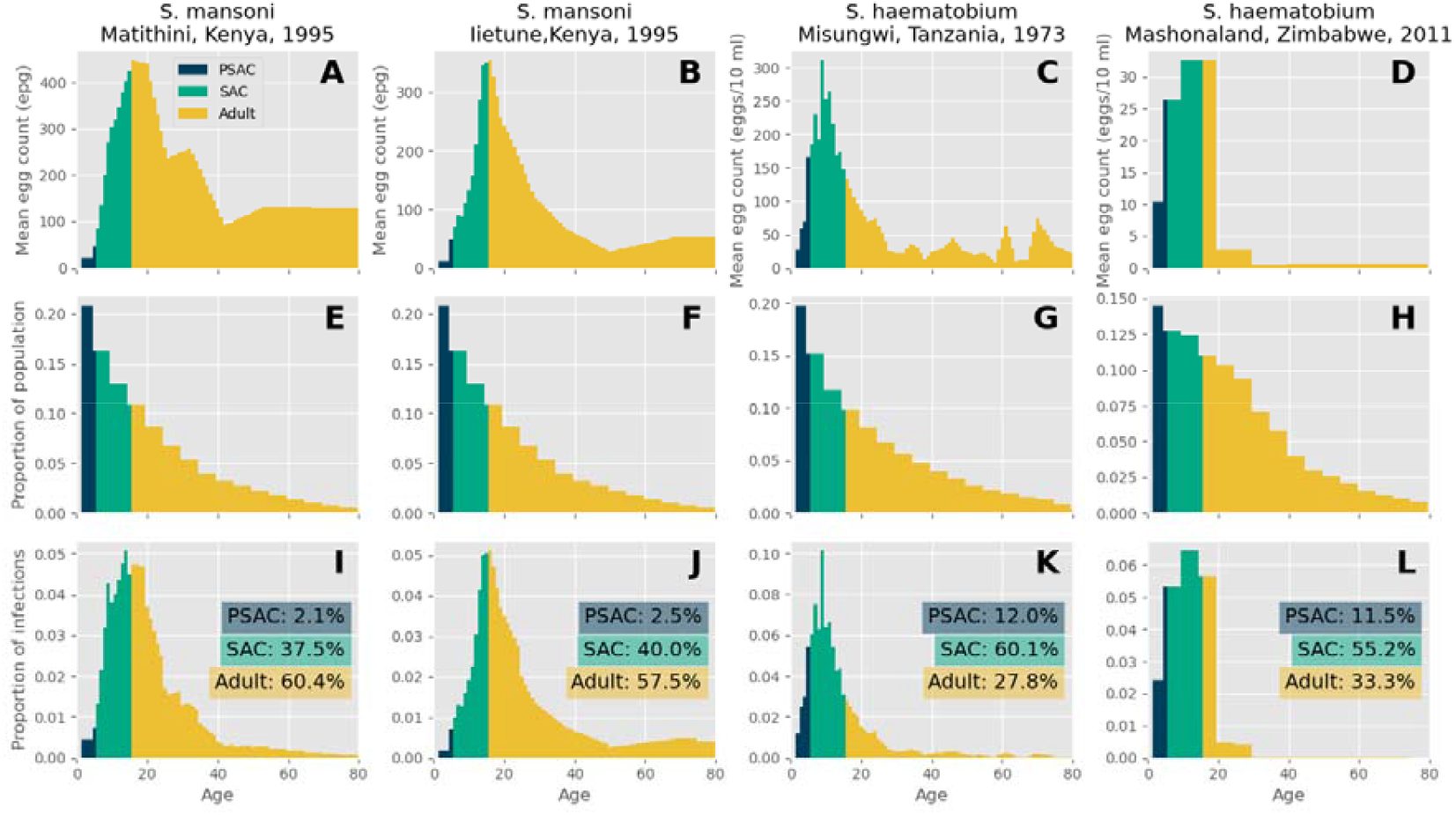
Four examples of the mean egg count (A-D), age distribution (E-H) and proportion of all infections (I-L) from previously published surveys. Panels A and B show mean egg counts of S. mansoni from two 1995 surveys in Matithini and Iietune, Kenya, respectively.^25^ Panel C shows the mean egg count of S. haematobium from a survey in Misungwi, Tanzania,^24^ and Panel D shows the mean egg count of S. haematobium from a 2011 survey in Mashonaland, Zimbabwe.^23^ Panels E-H show the corresponding national demography taken from the closest available year to the survey in the above panel in the US Census Bureau International Database.^26^ Panels I-L show the distribution of all infections in a community assuming the distribution of infection intensity with age and the demography in the panels within the same column, with the results broken down into PSAC, SAC and adults.

### Progress towards achieving the WHO goals for schistosomiasis

For the age profiles given in the Tanzania, and Matithini, Kenya datasets, which we will henceforth refer to as Settings One and Two respectively, we simulated the effect of targeting different combinations of age groups during MDA given the modern-day demography of Tanzania. Table 1 shows the probability of achieving EPHP, defined as <1% of heavy-intensity infection in SAC,^2^ after 5, 10, 15 and 20 years of MDA while varying the age groups that are treated. Coverage of all treated age groups is either 60% or 75%.

**Table 1:** The probability of achieving EPHP through MDA when treating different combinations of age groups with a 60% or 75% coverage. Results are calculated from 300 simulation runs of the model in Settings One and Two.

|  |  | 60% coverage |  |  |  | 75% coverage |  |  |  |
| --- | --- | --- | --- | --- | --- | --- | --- | --- | --- |
|  |  | SAC | PSAC and SAC | SAC and adult | All groups | SAC | PSAC and SAC | SAC and adult | All groups |
|  |  | Setting One: *S. haematobium*, Misungwi, Tanzania.<sup>24</sup> |  |  |  |  |  |  |  |
| Years of MDA | 5 | 0.3% | 0.3% | 0.0% | 0.0% | 1.0% | 4.0% | 1.7% | 9.3% |
|  | 10 | 5.7% | 13.7% | 9.0% | 25.0% | 45.0% | 69.7% | 64.3% | 87.7% |
|  | 15 | 39.0% | 57.3% | 45.7% | 72.7% | 92.3% | 96.7% | 96.7% | 99.7% |
|  | 20 | 70.7% | 85.0% | 80.0% | 92.7% | 100.0% | 100.0% | 100.0% | 100.0% |
|  |  | Setting Two: *S. mansoni*, Matithini, Kenya.<sup>25</sup> |  |  |  |  |  |  |  |
| Years of MDA | 5 | 0.0% | 1.0% | 18.7% | 17.7% | 2.3% | 3.3% | 69.0% | 65.3% |
|  | 10 | 4.3% | 5.0% | 84.3% | 82.7% | 15.3% | 18.0% | 100.0% | 99.3% |
|  | 15 | 13.0% | 14.3% | 98.3% | 99.3% | 36.3% | 44.3% | 100.0% | 100.0% |
|  | 20 | 24.0% | 25.7% | 100.0% | 100.0% | 61.3% | 62.0% | 100.0% | 100.0% |

The chances of EPHP varies greatly between the two settings, even when the same strategy is used. For example, if there is 60% coverage, in Setting One there is only a 90%+ chance of EPHP when all groups are treated for 20 years, whereas this can be achieved in Setting Two when SAC and adults are treated for 15 years. However, if adults are not treated, there is still a 90%+ probability of EPHP if other groups are treated with 75% coverage, but that is not possible in Setting Two. Treatment of PSAC does increase the probability of EPHP compared to treatment of SAC alone in Setting One, particularly when coverage is low (39% to 57.3% after 15 years at 60% coverage), or after fewer rounds of MDA (45% to 69.7% after 10 years at 75% coverage), however the increase is almost negligible in Scenario Two.

### Childhood morbidity

For morbidity, measured by WY, results are shown in Table 2. This reveals that it is more effective to achieve a 60% coverage of PSAC and SAC than a 75% of SAC alone in the first five years of MDA. However, this relationship reverses after 10 years, as a 75% coverage of SAC reduces the overall levels of transmission more effectively and can achieve EPHP, as shown in Table 1. Despite only considering WY for children aged five, in Setting Two it is more effective to treat adults alongside SAC than it is to treat PSAC.

**Table 2:** The reduction in worm years of five-year-olds during MDA when treating different combinations of age groups with a 60% or 75% coverage. Results are calculated using the mean WYs from 300 simulation runs of the model in Settings One and Two.

|  |  | 60% coverage |  |  |  | 75% coverage |  |  |  |
| --- | --- | --- | --- | --- | --- | --- | --- | --- | --- |
|  |  | SAC | PSAC and SAC | SAC and adult | All groups | SAC | PSAC and SAC | SAC and adult | All groups |
|  |  | Setting One: *S. haematobium*, Misungwi, Tanzania.<sup>24</sup> |  |  |  |  |  |  |  |
| Years of MDA | 5 | 43.8% | 68.4% | 47.3% | 69.5% | 54.1% | 75.8% | 57.4% | 77.5% |
|  | 10 | 83.3% | 91.8% | 84.0% | 93.6% | 91.8% | 96.8% | 94.4% | 98.3% |
|  | 15 | 93.4% | 97.2% | 93.5% | 98.3% | 98.4% | 99.5% | 99.1% | 99.9% |
|  | 20 | 97.3% | 99.1% | 98.2% | 99.5% | 99.7% | 99.9% | 99.9% | 100.0% |
|  |  | Setting Two: *S. mansoni*, Matithini, Kenya.<sup>25</sup> |  |  |  |  |  |  |  |
| Years of MDA | 5 | 37.7% | 52.8% | 63.4% | 74.9% | 42.6% | 58.1% | 70.8% | 83.7% |
|  | 10 | 58.5% | 68.9% | 92.0% | 94.3% | 66.8% | 76.6% | 97.5% | 98.6% |
|  | 15 | 69.4% | 76.8% | 97.7% | 98.3% | 79.1% | 85.3% | 99.7% | 99.9% |
|  | 20 | 75.7% | 81.9% | 99.4% | 99.7% | 85.4% | 89.5% | 100.0% | 100.0% |

### Variation in the age profile of the intensity of infection and host demography

The impact of variations in the age intensity of infection profile are illustrated in Figure 2A, where an increase in β_0-5_, the parameter controlling the relative contact rate in the 0-5 age group (see Table S1 in the supplementary material), leads to an increase in the mean egg count in ages 0-10. This leads to an increase in the POI found in PSAC, as shown in Figure 2B, ranging from 1.6% to 21.1% while there is a slight decrease in the POI in SAC and adults, from 73.6% and 24.7%, to 59.4% and 15.7% respectively.

**Figure 2:**
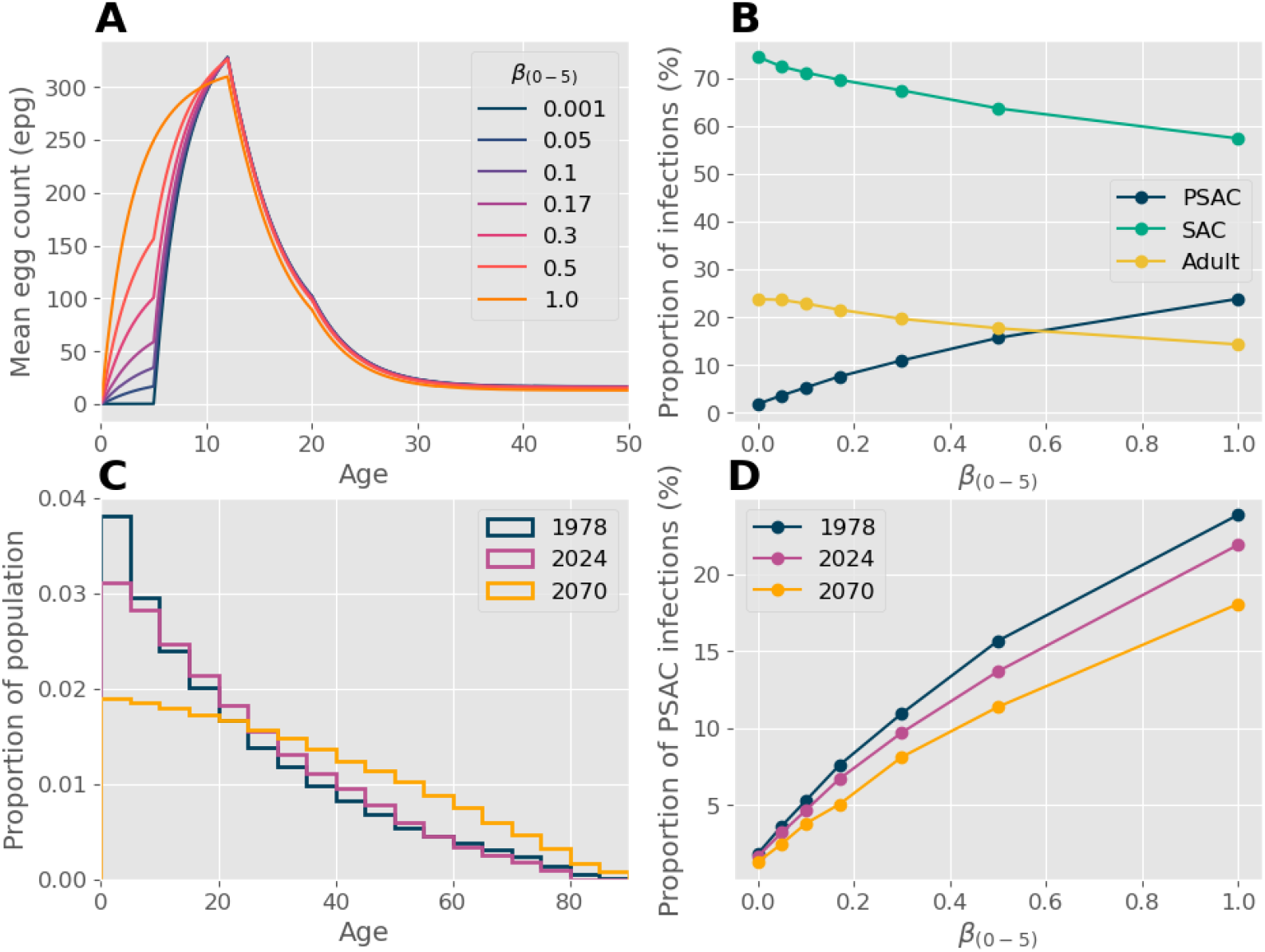
Variation in the proportion of infections when changing the relative contact rates for ages 0-5 and the demography. Panel A shows the infection intensity with age, used in the model with β_(0-5)_ ranging from 0.001 (low probability of infection) to 1 (high probability of infection). Panel B shows the resulting proportion of infections in each age group when changing β_(0-5)_. Panel C shows the estimated/projected demography of Tanzania in 1978, 2024 and 2070 and Panel D shows the resulting change in the proportion of infections in PSAC due to the change in demography.

The Host demographic age profile impacts the POI by changing the proportion of the population in each age group. Figure 2C shows the demography for Tanzania years: 1978, 2024 and a projection for 2070. The population is concentrated in younger age groups in 1978, but by 2070 the age profile is predicted to be much flatter, with a higher proportion of adults in the population. The implications for this are that in 2070, the POI in PSAC will be less than in 1978 and 2024, as shown in Figure 2D, with bigger differences in areas which have a high infection rate in PSAC. For the results shown below, we use the 2024 demography. Results for 1979 and 2070 are recorded in the supplementary material, Figs S1 and S2.

The result of an increase in POI in PSAC is a reduction in the impact of MDA when PSAC are not treated, as shown by the dark lines in Figure 3A and B. When the POI in PSAC at baseline increases, the probability of reaching EPHP by 5 or 10 years drops significantly. When the POI in PSAC is low, treating PSAC makes no difference to the probability of EPHP. However, for high PSAC infections, the difference can be substantial, particularly when there is the potential for a high probability of EPHP, as is the case after 10 years with 75% SAC and adult coverage. If coverage in other age groups is low, treating PSAC can increase the probability of reaching EPHP when the POI in PSAC is high (>20%), but only to approximately 35% compared to 80% when the coverage is 75% in SAC and adults.

**Figure 3:**
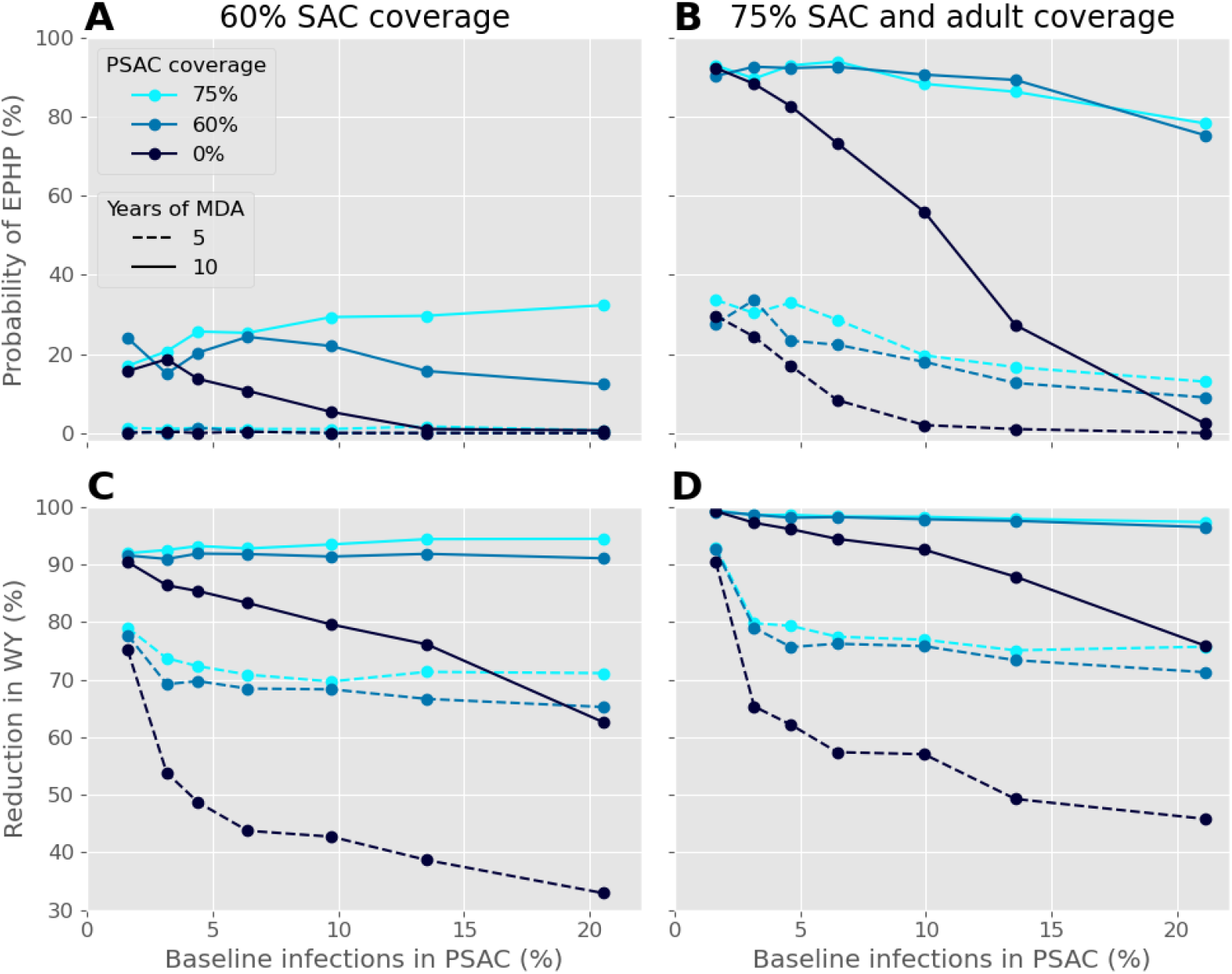
The probability of EPHP (Panels A and B) and the reduction in WY (Panels C and D) after 5 and 10 years of MDA with 0, 60 or 75 coverage of PSAC when varying the proportion of infections in PSAC at baseline. In Panels A and C, coverage in other age groups is limited to just 60% of SAC and in Panels B and D, coverage is 75% for both SAC and adults.

Coverage in other age groups is less important when examining the relationship between POI in PSAC and the reduction in WY for 5-year-olds, as shown in Figure 3C and D. When the POI in PSAC is above 2%, the effect from increasing PSAC coverage is much more than increasing coverage in other age groups. Comparing individual points in the plots shows a maximum difference of 15% when switching from 60% SAC coverage to 75% SAC and adult coverage. Whereas, after 5 years the difference between treating or not treating PSAC is substantial, ranging from 20% to 40%. After 10 years the difference is reduced; with treatment of PSAC, the reduction in WY approaches 100%, but without treatment of PSAC, the impact is dependent on the POI in PSAC. Note that for higher POI in PSAC, it follows that the average WY will be higher before treatment, meaning that the percentage reduction in WY will translate to a higher absolute reduction than at a lower POI in PSAC.

## Discussion

Our results demonstrate that the proportion of the total worm population (POI) in PSAC prior to widespread MDA is a key factor in determining the effectiveness of treating that age group with the novel praziquantel formulation for young children. A high POI in PSAC will increase the necessity of including PSAC in MDA. However, if the POI in PSAC is low, there may be little benefit when only considering the goal of EPHP or other prevalence reduction targets. If additionally, the POI in adults is high, then including adults in MDA may be more beneficial to PSAC than treating PSAC. This seems counterintuitive at first sight. The explanation lies in the reduction in infections amongst adults which will, concomitantly, reduce the transmission rate in all age groups and will therefore benefit those that might not have attended for treatment (they acquire indirect benefits).

Infection levels in PSAC is receiving much more attention in recent years, due to the recognition of the need for treatment in young children to prevent future morbidity induced by early infection. The potential availability of L-PZQ has been an added stimulus. However, it remains true that most of the epidemiological surveys conducted in country-based monitoring and evaluation surveys to assess progress in control rarely considers all age groups in a population.^30^ They usually focusing on SAC, or PSAC and their mothers.^8,9^ Knowledge of the distribution of infection across all ages is required to calculate the POI in PSAC. As illustrated in the simulations in this paper, this detail matters in determining the potential impact of including PSAC in MDA. Where data across all ages is available, studies show a range of patterns in the age distribution of infection intensity, particularly in PSAC. The age distribution may also vary greatly even in between villages in a defined locality.

The MDA strategy that is the best at achieving EPHP, will not necessarily be the best at reducing childhood morbidity as measured by the WY a child experiences. IF EPHP is achievable, then it will clearly have benefits for the whole population. However, if EPHP cannot be achieved in a time span of say 5-10 years of MDA, either because coverage is not adequate or there are other issues not considered in our model such as the continued outside introduction of infection due to people movements or migration or zoonotic transmission, then the simulations suggest that including PSAC in MDA will be the best method of reducing morbidity long term.

We do not consider the costs of treatment in this study; however, we note that there will be costs associated with manufacturing and distributing the new drug formulation. As yet these are not clearly defined. In practice on the ground in resource poor settings t may be easier to achieve high coverage of PSAC as they make up a smaller proportion of the population than adults and may be much easier to locate for treatment. As has been shown previously, a higher burden of infection in adults makes it more difficult to achieve control targets often as a result of low MDA coverage levels in this age grouping.^21^ In these circumstances, it is more worthwhile to expend resources to include the adult population in MDA, while treatment of PSAC could occur on a case-by-case basis. A full economic analysis when costs of PSAC treatment are better understood is highly desirable.

A key objective of this study is to demonstrate the importance of understanding the full distribution of infections by age in the design of control programmes for schistosomiasis. In areas with a low PSAC burden of infection, decision makers should not expect that introducing PSAC into MDA will greatly increase the chances of achieving EPH. When the PSAC burden is high prior to MDA, elimination as a public health problem will be difficult to achieve unless high coverage of SAC is also attained. Instead, a successful implementation of MDA to PSAC should be regarded as one that improves childhood health, where more immediate benefits will be achieved in terms of reducing ‘wormy years’ for children. Although a metric such as WY is not measurable outside of simulations, reduction in stunting rates and improvements in school attendance would be possible measurable outcomes in longitudinal epidemiological studies of control impact.

## Data Availability

All data produced in the present work are contained or referenced in the manuscript.

