## supplementary material for "Pre-school Age Participation in Mass Drug Administration: Analysing the Impact on Community-wide Schistosomiasis Control"

Supplementary Materials

Table S1: Parameter values for Schistosoma haematobium used for Setting One.

| **Parameter** | **Value** | **Reference** |
| --- | --- | --- |
| Fecundity (eggs/female/10 ml sample) | 3.6 | [1], [2] |
| Aggregation parameter | 0.24 | [3] |
| Density dependent fecundity | 0.0006 | [4] |
| Worm life span (years) | 4 | [5], [6] |
| Age specific contact rates for 0-5, 5-12, 12-20, 20+ years old | 0.17, 1, 0.11, 0.035 | [7] |
| Drug efficacy | 94% | [8] |
| Aggregation of diagnostic | 0.5 | [3] |
| Basic reproduction number | 2.0 | - |
| Population size | 500 | - |

Table S2: Parameter values for Schistosoma mansoni used for Setting Two.

| **Parameter** | **Value** | **Reference** |
| --- | --- | --- |
| Fecundity (eggs/female/10 ml sample) | 3.6 | [1], [2] |
| Aggregation parameter | 0.24 | [3], [4] |
| Density dependent fecundity | 0.0007 | [4], [9] |
| Worm life span (years) | 5.7 | [4], [10], [11] |
| Age specific contact rates for 0-5, 5-12, 12-20, 20+ years old | 0.01, 0.61, 1, 0.12 | [9], [12] |
| Drug efficacy | 86.3% | [8] |
| Aggregation of diagnostic | 0.87 | [13], [14], [15] |
| Basic reproduction number | 2.0 | - |
| Population size | 500 | - |

### Setting Two: *S. mansoni*

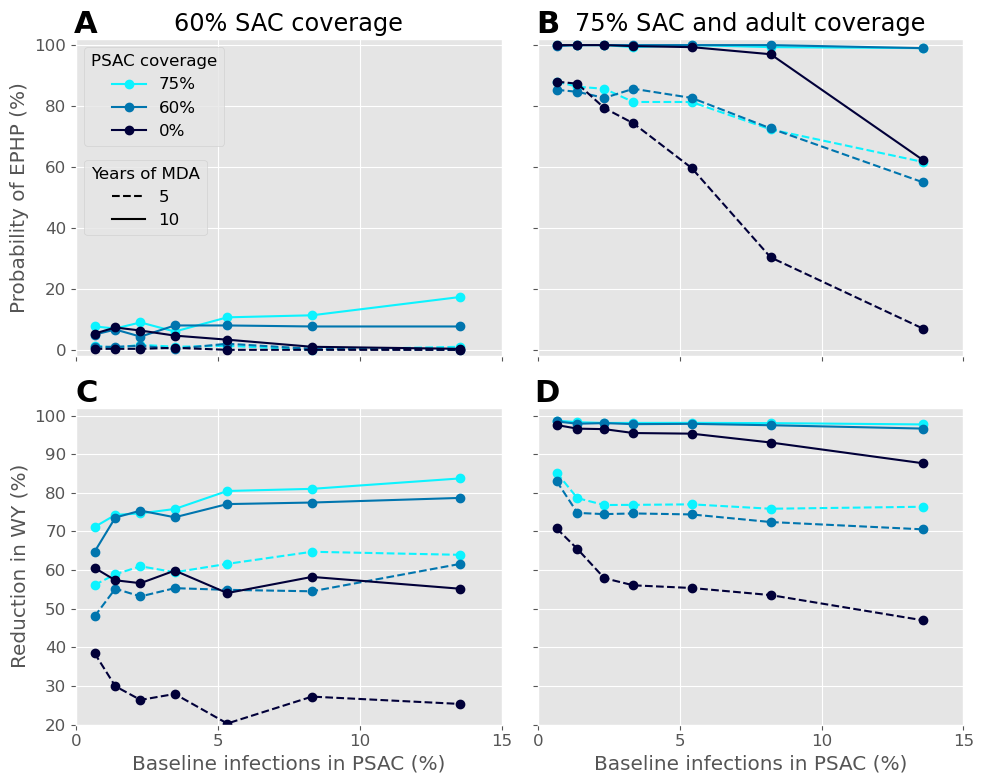

Figure S1: Results of the model of S. mansoni in Setting Two. The probability of EPHP (Panels A and B) and the reduction in WY (Panels C and D) after 5 and 10 years of MDA with 0, 60 or 75 coverage of PSAC when varying the proportion of infections in PSAC at baseline. In Panels A and C, coverage in other age groups is limited to just 60% of SAC and in Panels B and D, coverage is 75% for both SAC and adults.

### Changes in demography

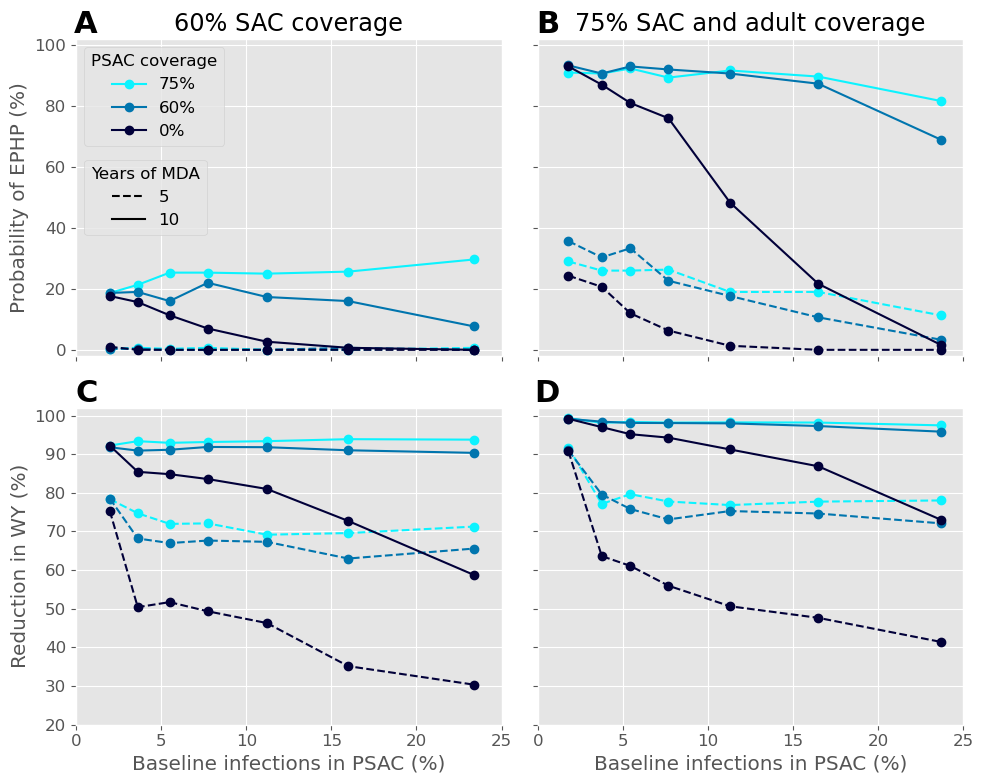

Figure 2: Results of the model using demography from 1978. The probability of EPHP (Panels A and B) and the reduction in WY (Panels C and D) after 5 and 10 years of MDA with 0, 60 or 75 coverage of PSAC when varying the proportion of infections in PSAC at baseline. In Panels A and C, coverage in other age groups is limited to just 60% of SAC and in Panels B and D, coverage is 75% for both SAC and adults.

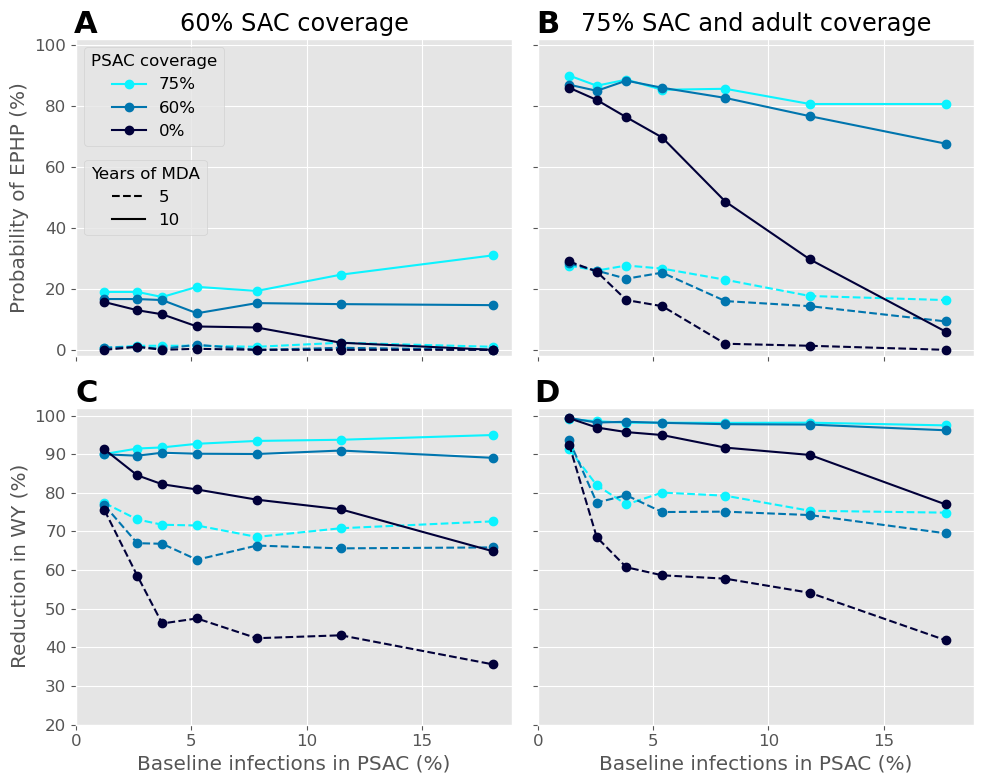

Figure 3: Results of the model using the projected demography from 2070. The probability of EPHP (Panels A and B) and the reduction in WY (Panels C and D) after 5 and 10 years of MDA with 0, 60 or 75 coverage of PSAC when varying the proportion of infections in PSAC at baseline. In Panels A and C, coverage in other age groups is limited to just 60% of SAC and in Panels B and D, coverage is 75% for both SAC and adults.
